# Addressing algorithmic bias in precision well-being for medical education: A socially fair approach for clustering

**DOI:** 10.1101/2024.12.10.24318825

**Authors:** Priyanshu Alluri, Zequn Chen, Wesley J. Marrero, Nicholas C. Jacobson, Thomas Thesen

## Abstract

**Background:** Medical students frequently experience heightened levels of anxiety, depression, and burnout. These challenges are disproportionately borne by students from underrepresented backgrounds, who are exposed to systemic inequities, discrimination, and reduced access to supportive resources. While precision well-being approaches, characterized by identifying distinct well-being phenotypes for personalized interventions, hold promise, standard machine learning clustering algorithms such as K-Means may inadvertently exacerbate these disparities. Furthermore, the underlying factors contributing to poorer mental health outcomes among underrepresented students remain insufficiently understood.

**Objective:** We aim to identify well-being phenotypes that achieve an equitable distribution of clustering costs across racial groups, identify conditions under which fair and standard clustering solutions converge, and investigate the demographic and socioeconomic factors that shape mental health patterns in students underrepresented in medicine.

**Methods:** Drawing on a diverse sample of 4161 medical students from multiple U.S. institutions participating in the Healthy Minds Survey (2016–2021), we compared the outcomes of socially fair and standard k-Means clustering algorithms using Patient Health Questionnaire-9, General Anxiety Disorder-7, and Flourishing scores. We then employed average treatment effect analyses to identify factors that exacerbate mental health challenges and those that enhance resilience, with a particular emphasis on underrepresented populations.

**Results:** The socially fair clustering algorithm significantly reduced the disproportionate burden on minority populations, aligning with standard clustering outcomes when student groups were racially and socioeconomically homogeneous. Perceived discrimination emerged as a key factor driving poorer mental health, while stable financial conditions, robust social engagement, and involvement in culturally or ethnically oriented organizations were linked to greater resilience and improved well-being.

**Conclusions:** Incorporating fairness objectives into clustering algorithms substantially reduced the disproportionate burden on minority students and yielded a more equitable understanding of their mental health patterns. By identifying factors that influence mental health outcomes, our socially-fair precision well-being approach allows for more personalized well-being interventions. These insights equip educators and policymakers with actionable targets for developing culturally responsive, data-driven interventions that not only alleviate distress but also support resilience, ultimately advancing more inclusive, effective precision well-being strategies for all medical students.

## Background

Medical students are particularly vulnerable to mental health disorders due to the intense pressures of their training (1,2). Studies consistently show medical students experience high rates of anxiety, depression, and burnout, significantly impacting their well-being and academic performance (3–6). This issue is especially pronounced among minority students, who often face particular challenges such as systemic discrimination (7,8), lack of representation (9,10), and social isolation (4), leading to a disproportionate burden of mental health challenges (11–13). Although the factors contributing to this increased burden in underrepresented medical students are likely multifactorial, ranging from structural inequities to cultural, financial, and interpersonal stressors, their precise roles and interactions remain insufficiently defined in the existing literature (14,15).

To better address the mental health needs of medical students, healthcare professionals often identify distinct subgroups of students with similar mental health profiles using psychological and behavioral indicators (16,17). For instance, a recent study using K-Means clustering on data from over 3,600 medical students identified three main clusters: “Healthy Flourishers,” “Getting By,” and “At-Risk” (18). By revealing patterns in symptoms, coping strategies, and levels of psychological distress, such clustering approaches create well-being phenotypes that can help guide more personalized, evidence-based interventions, ensuring that those at greatest risk receive the targeted resources and support they need.

However, traditional clustering algorithms, such as the standard K-Means, often result in biased outcomes that disproportionately assign a higher cost burden to minority populations (19–21). Standard K-Means focuses solely on minimizing overall differences within clusters, without considering how those differences are spread across different demographic groups. This approach may amplify structural inequalities by assigning an outsized within-cluster variance to underrepresented groups, reducing the accuracy of the grouping of minority populations. An inequitable distribution of variances may unintentionally group minority students in ways that reflect and reinforce existing inequalities, rather than offering an accurate understanding of their mental health needs. As a result, standard K-Means clustering can lead to less effective interventions that exacerbate existing disparities between racial groups (22).

Fair clustering algorithms have been developed to address these inequities and ensure a more equitable distribution of clustering costs across demographic groups (23). Socially Fair Clustering, a promising approach in the fair clustering domain, modifies traditional algorithms by incorporating a fairness objective, thereby creating clusters that are both reflective of the data structure and equitable toward minorities (21). This approach is especially relevant in mental health clustering, where it is vital to avoid further marginalizing vulnerable populations such as racial and ethnic minority medical students.

In this study, we consider fairness with respect to race and apply the socially fair K-Means clustering algorithm to medical student mental health data. We then compare the clustering costs for minority populations under socially fair clustering and standard K-Means clustering. Beyond race, we also examine the influence of various demographic and socioeconomic factors on clustering outcomes.

## Methods

### Data Preparation

The data for this study is sourced from the Healthy Minds Survey, provided by the Healthy Minds Network. This dataset comprises survey responses from a diverse sample of medical students across the United States, collected between January 1, 2016, and December 31, 2021. The survey encompasses demographic and socioeconomic characteristics, mental health status, mental health service utilization, and related factors (17).

Survey respondents aged 18 and older who answered the Patient Health Questionnaire-9 (PHQ-9) (24), General Anxiety Disorder-7 (GAD-7) (25), and Flourishing Scale (26) questionnaires are selected. These metrics are chosen because they are widely used for screening mental health issues (27–29), have been clinically validated (24–26,30), and have been used in previous studies with similar applications (18, 31, 32). We exclude responses missing race data, PHQ-9, GAD-7, or flourishing scale items from the analysis to avoid statistical uncertainty in our outcomes and sensitive attributes.

To create composite scores, all PHQ-9 sub-questions are summed to generate a single PHQ-9 score, and this process is repeated for the GAD-7 and flourishing scale items. Race responses are converted to a single categorical variable with values corresponding to White, Asian, Black, Hispanic, and Other. Respondents identifying as Middle Eastern, Pacific Islander, American Indian, other, or mixed are grouped under the “Other” category.

These three combined variables (PHQ-9, GAD-7, flourishing) are normalized to a range of 0 to 1. Race labels for each sample are maintained, and all other variables in the dataset are excluded from the clustering procedures.

### Standard Clustering

The K-Means clustering algorithm is widely used to partition a dataset into *k* distinct clusters, minimizing the total within-cluster variance. We choose K-Means because it provides a scalable, efficient, and interpretable approach to clustering. Given our data’s characteristics and the need for clear, well-defined clusters, K-Means is ideal for efficiently delivering reliable and meaningful clusters. Given an input of points *U*, the K-Means algorithm assigns each data point in *U* to the nearest cluster center by Euclidean distance (22). Each cluster center is then recalculated as the average of all points in that cluster. This process is repeated until convergence (33). The K-Means clustering algorithm minimizes the total within-cluster variance, defined as:

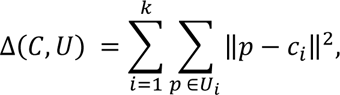

where *C* denotes the set of all cluster centers, *U* denotes the set of all points to be clustered, *U_i_* represents the subset of points in *U* that are assigned to an individual cluster center *c_i_*, and *p* denotes an individual data point.

### Socially Fair Clustering

The standard K-Means clustering algorithm often results in biased clusterings that can disproportionately affect minority racial groups (34). In this study, we mitigate the effects of this burden using the fair K-means algorithm (21). This algorithm modifies the traditional K-Means heuristic to minimize the *highest* within-cluster variance cost across two different demographic groups, *A* and *B*:

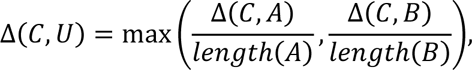

where *length* is the number of individuals in each group (21). In this setting, higher costs indicate greater intra-group dissimilarity.

The fair K-means algorithm operates given an input of points *U*, such that all points in *U* belong to either group *A* or group *B*. A set of centers is initialized by randomly selecting *k* data points from the dataset *U*. Each data point in *U* is assigned to the nearest center to form *k* clusters. The *k* centers are then updated to minimize the maximum average clustering cost across different demographic groups. Each data point in *U* is then reassigned to the updated *k* cluster centers. This process is repeated until convergence.

We generate fair clusterings for each race pair (White-Black, White-Hispanic, White-Other, etc.). These clusterings are then compared with those observed for each race pair without applying the social fairness objective.

### Clustering Evaluation

To assess our clustering results, we bootstrap the data from each race. The size of each bootstrap resample corresponds to the race with the smallest number of observations, and 10,000 bootstrap resamples are generated for each race (35). For each resample, we calculate the average clustering cost for each race (36). We repeat this process for each fair K-means clustering and each standard K-Means clustering.

### Statistical Analysis of Clusterings

To explain the behavior of different racial groups, we first add all variables recorded in the Healthy Minds Survey to the analysis. This includes responses to demographic questions, lifestyle questions, and other miscellaneous questions. We thus add a total of 3227 variables to the analysis. We then use analysis of variance to identify all numeric variables that have a statistically different mean value between the groupings (37). The statistically significant variables are analyzed using Tukey’s Honestly Significant Difference test as a post-hoc analysis to identify which specific groups differ from each other (38).

The chi-square test of independence is employed to determine whether there is a significant association between race and binary variables in the dataset. The statistically significant variables are analyzed using a post-hoc chi-square test with the Bonferroni correction to compare the proportions between specific racial groups. The Kruskal-Wallis H test is employed to assess whether the distributions of ordinal variables differed significantly between racial groups. The statistically significant variables are then analyzed using Dunn’s test as a post-hoc analysis to determine the specific group differences.

Significant variables identified through a primary significance test and post-hoc analysis are appended to each data point used in clustering. For each cluster (“Flourishing,” “Getting By,” and “At Risk”), the 20th and 80th percentile values of each variable are calculated, excluding any missing values. This process is repeated for both the standard K-Means clusterings and the fair K-Means clusterings. To identify influential variables, the range between the 20th and 80th percentiles for each variable is compared across the “Flourishing” and “At Risk” clusters. Variables with non-overlapping ranges between these clusters are flagged as potentially influential and selected for further analysis.

The analyses are conducted in Python, utilizing the scipy.stats package (39).

### Average Treatment Effect

We employ the Average Treatment Effect (ATE) metric to determine whether there is evidence that other variables in the dataset influence changes in PHQ-9, GAD-7, and flourishing scores. This metric represents the mean outcome difference between treated and control groups across a population. One robust method for estimating ATE is Double Machine Learning (DML), which leverages machine learning models to control for confounding variables (40). We employ the econml library’s DML method to estimate the ATE of variables with distinct separation by cluster on PHQ-9, GAD-7, and flourishing scores (41).

Separate linear regression models are specified for each of the scores. A logistic regression model is specified for each treatment variable. When the treatment variable is binary, one condition is selected as the treatment, and the other as the control. For ordinal variables with more than two possible answers, a threshold is set where the absolute difference in treatment effect is maximized. Values above the threshold are considered the treatment condition, and values below the threshold are considered the control condition.

The LinearDML estimator is then instantiated, recognizing that each treatment variable is discrete. The linear version of the DML estimator is chosen over other options because it works best when the number of values a feature can take is significantly less than the number of samples, as in our dataset. The ATEs are calculated separately for the flourishing, PHQ-9, and GAD-7 scores. Variables are then ranked based on the absolute value of their ATEs on flourishing scores, PHQ-9 scores, and GAD-7 scores.

### Practice-Inspired Clustering Solutions

Practitioners may benefit from having access to a clustering solution with three distinct centers, to which they can provide tiered treatments (42). We therefore modify the standard and fair K-Means clustering algorithms to choose the minimum number of centers, *k*, that achieve three distinct subgroups. Three centers corresponding to flourishing, getting by, and at risk clusters are initialized by selecting one representative center from the list of *k*. Every other center is then assigned to the flourishing, getting by, or at-risk subgroup by euclidean distance. The centers from each subgroup are then averaged to generate a three-center solution. Using these centers, each data point is assigned to the nearest cluster.

## Results

In this section, we present our results on the application of the socially fair K-Means algorithm for clustering medical students based on mental health groupings. We explore how this algorithm compares to traditional K-Means clustering and highlight key insights from the data.

Clustering costs were quantified by calculating within-cluster variance for each group, with higher costs indicating greater intra-group dissimilarity. We find that even when fairness is prioritized, the “Black” and “Other” racial groups incur higher clustering costs than other groups. However, the socially fair K-Means algorithm still leads to a considerable reduction in clustering costs for minority populations when compared to the standard K-Means approach. We also observe that the fair clustering algorithm tends toward a two-center solution rather than the three-center solution from the standard K-Means in prior work (18). Furthermore, our findings suggest that when racial groups tend to experience similar levels of fear about discrimination, the non-fair clustering solution approximates the fair clustering solution.

These findings emphasize the importance of incorporating fairness objectives in clustering algorithms, especially when working with minority groups that typically bear higher clustering costs in traditional approaches.

### Clustering Costs Across Racial Groups

Our analysis begins with a comparison of clustering costs between the socially fair and standard K-Means algorithms. As expected, the standard K-Means algorithm results in higher clustering costs for minority populations. The fair clustering algorithm can only accommodate two race groups to generate fair centers, so we generate standard cluster centers for each racial pair and conduct 10,000 bootstrap resamples of size 192 for each group. A bootstrap resample size of 192 students is chosen because the race with the smallest size, “Other,” had 192 students. These resamples are used to calculate the average clustering cost for each group (Fig. 1, S1, and S2).

**Figure 1.**
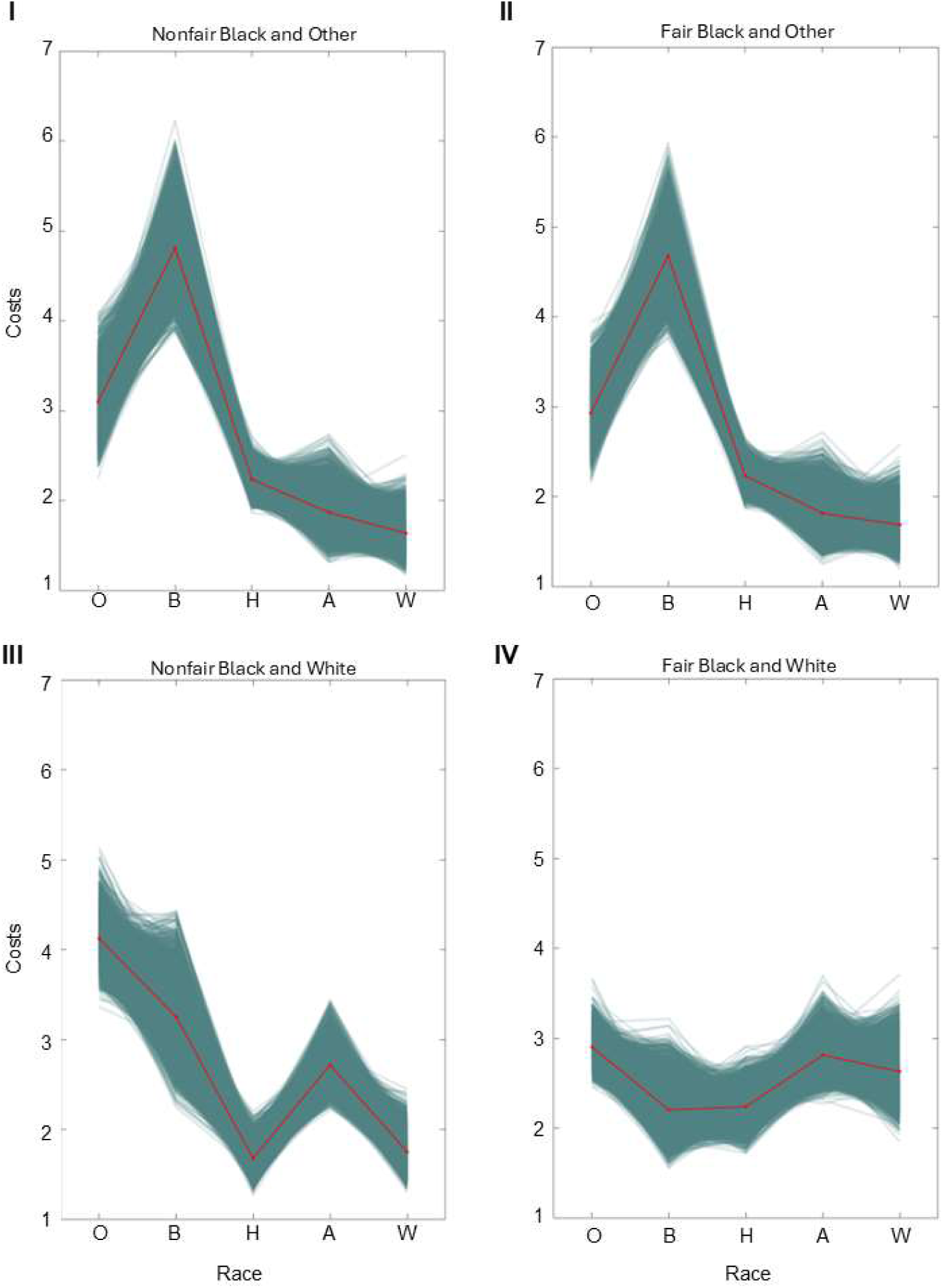
Average clustering costs across Other (O), Black (B), Hispanic (H), Asian (A), and White (W) races from 10,000 bootstrap samples when clustering Black and Other populations in the standard K-Means algorithm (I), and in the socially fair K-Means algorithm (II). Average clustering costs across races from 10,000 bootstrap samples when clustering Black and Other populations in the standard K-Means algorithm (III), and in the socially fair K-Means algorithm (IV).

The results consistently demonstrate that “Black” and “Other” racial minority groups experience the highest clustering costs across all non-fair clusters (Fig. 1I and 1III). When we apply the fair K-means algorithm, these costs are drastically reduced and more equitably distributed across all racial groups, although “Black” and “Other” groups still have higher costs in most clusterings (Fig. 1II, 1IV, and S2). This finding underscores the need for fairness algorithms to help reduce disparities, though further refinements may be necessary to fully eliminate inequities.

Additionally, the fair and standard K-Means clustering of groups with similar socioeconomic status, such as the clustering of Black and Other groups, results in only minimal differences in cost, while the fairness objective significantly lowers minority costs when clustering groups of differing socioeconomic status (Fig. S1 and S2).

To understand why the “Black” and “Other” groups consistently bear the highest costs, we further examine the within-group variance in PHQ-9, GAD-7, and flourishing scores. The analysis reveals that “Black” and “Other” groups have the highest variances, which likely partially contributes to their high clustering costs (Fig. S3). This finding underscores the importance of accounting for within-group variance when evaluating clustering outcomes.

### Fair Clustering Solution’s Convergence to a Two-Center Model

The standard K-Means algorithm previously identified three clusters: one for students at risk, one for those getting by, and one for those flourishing (18). In contrast, the socially fair K-Means algorithm consistently converges to a two-center solution, distinguishing between students at risk and those flourishing (Fig. 2 and S4).

**Figure 2.**
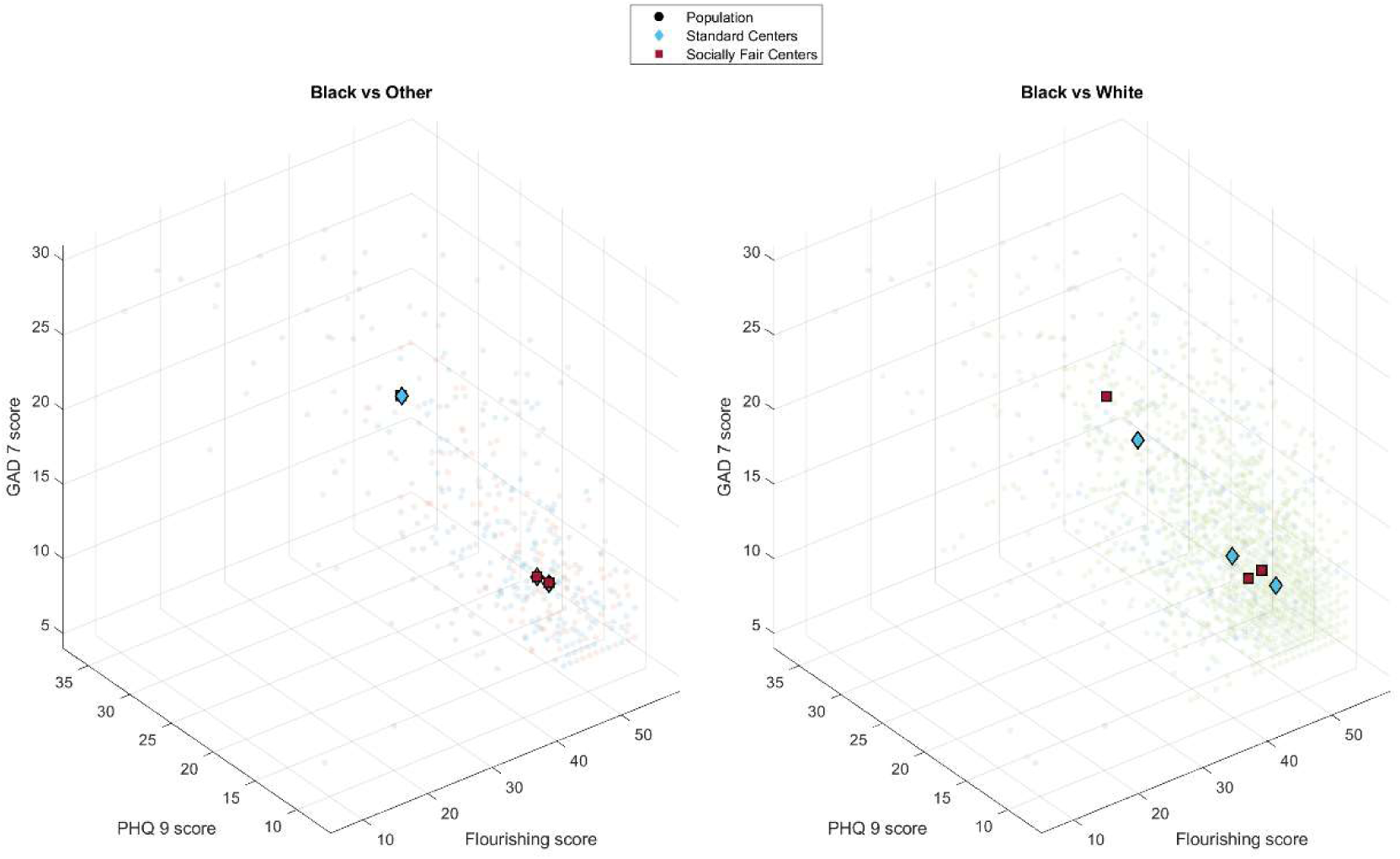
(Left) Overlay of cluster centers generated by the socially fair K-Means algorithm and standard K-Means algorithm when clustering “Black” and “Other” racial groups. (Right) Overlay of cluster centers generated by the socially fair K-Means algorithm and standard K-Means algorithm when clustering “Black” and “White” racial groups.

Analysis of the clustering costs identified that when paired with one another, Black, Hispanic, and Other groups showed little difference between the standard K-Means algorithm and the Socially Fair K-Means algorithm (Fig. S1 and S2). This suggested that Black, Hispanic, and Other student populations were sufficiently similar in their mental health profile that fairness constraints did not affect clusters much – they were already socially fair. Similarly, there was little difference between the standard K-Means algorithm and the Socially Fair K-Means algorithm when clustering Asian and White populations.

When the Black, Hispanic, or Other groups were clustered with Asian or White groups, however, there was a significant difference between the Standard K-Means algorithm and the Socially Fair K-Means Algorithm. This result indicates that the mental health profile of the Asian and White groups differs considerably from the Hispanic, Black, and Other group – fairness constraints are needed to equalize the cost across these disparate groups. Such a finding is also supported by the literature. Black, Hispanic, and Other groups are underrepresented in the medical student population, and they tend to face greater structural barriers and mental health challenges than their White and Asian peers (7,43,44).

Furthermore, we note that differences in clustering outcomes exist between Asian and White races as well. For instance, the Asian and Other clustering shows an equitable distribution of costs, while the White and Other clustering shows a disparate distribution of costs (Fig. S1). Since Asian medical students are known to have poorer outcomes than their white peers due to a combination of factors, including societal pressure and anxiety, we grouped Asian and white races as distinct groups (45,46).

Moreover, when clustering within one of these groups (Black, Hispanic, Other; Asian; White), such as in Fig. 2 left (Black vs Other), we note that the standard K-Means solution approaches the two-center socially fair K-means solution.

We therefore identified three groups: the underrepresented group, consisting of Black, Hispanic, and Other; the Asian race group; and the White race group. We then looked for variables that differed significantly between these three groups. Such variables could play a role in driving the differences in clustering results across groups and in explaining the mental health disparities between the groups.

### Statistical analysis of socioeconomic status factors

Each variable in the Healthy Minds Survey dataset was first analyzed to see if the variable differed significantly between any races. If this was the case, the variable was then tested with a post-hoc significance test to ensure that the variable had statistically significant differences between the three identified groups. Our statistical analyses of the socioeconomic factors contributing to differences in clustering results across the underrepresented, Asian, and White groups identify 17 variables that differ significantly between them. A portion of the 17 statistically significant variables are listed in Table 1; all 17 statistically significant variables are listed in Table S1. White students often have the most extreme scores, followed immediately by Asian students and then Black, Hispanic, and Other students in various orders.

**Table 1.** Select statistically significant variables that drive differences in clustering results across groups.

| Variable Name | Variable Question | Potential Options | Treatment |
| --- | --- | --- | --- |
| Current Financial Status | How would you describe your financial situation right now? | 1=Always stressful 2=Often stressful 3=Sometimes stressful 4=Rarely stressful 5=Never stressful | 4,5 |
| Past financial status | How would you describe your financial situation while growing up? | 1=Always stressful 2=Often stressful 3=Sometimes stressful 4=Rarely stressful 5=Never stressful | 4,5 |
| Active culturally | Are you involved in any cultural or racial organizations at your school? | 0=No 1=Yes | 1 |
| Parental birthplace | Were your parents born in the United States? | 1 = Yes 4 = No | 4 |
| Experience racial fear | In your classes, how often did you have fears of representing your racial/ethnic group in a negative way that discouraged you from participating in class? | 1=Almost never 2=Not very often 3=Sometimes 4=Fairly Often 5=Very Often | 2,3,4,5 |
| Discrimination experiences | In the past 12 months have you been treated unfairly at your school because of your race, culture, or gender? | 0 = No 1 = Yes | 1 |
| Fear of representing racial group | How often did you feel that others were taking your opinion as speaking for all members of your ethnic/racial group? | 1=Almost never 2=Not very often 3=Sometimes 4=Fairly Often 5=Very Often | 2,3,4,5 |
| Social involvement | How often do you attend meetings, events, activities, clubs, etc., that support your racial/ethnic identity? | 1 = Never 2 = Less than once a month 3 = 1 - 3 times a month 4 = Weekly 5 = Multiple times a week 6 = Every day | 4,5,6 |

The questions asked by the survey and the possible responses are listed for each variable. The responses treated as the treatment condition when conducting causal inference are listed in the "Treatment" column.

### Key Variables Influencing Clustering Patterns

To quantify the influence of the statistically significant variables (Table S1), we estimate their ATE on flourishing, PHQ-9, and GAD-7 scores. Variables that positively impact Flourishing scores and negatively affect PHQ-9 and GAD-7 scores are associated with better mental health outcomes. The variable with the greatest impact on mental health outcomes with respect to ATE is discrimination experiences, suggesting that perceived discrimination has the most substantial impact on mental health indicators, and thus clustering outcomes (Fig. 3 and S5).

**Figure 3.**
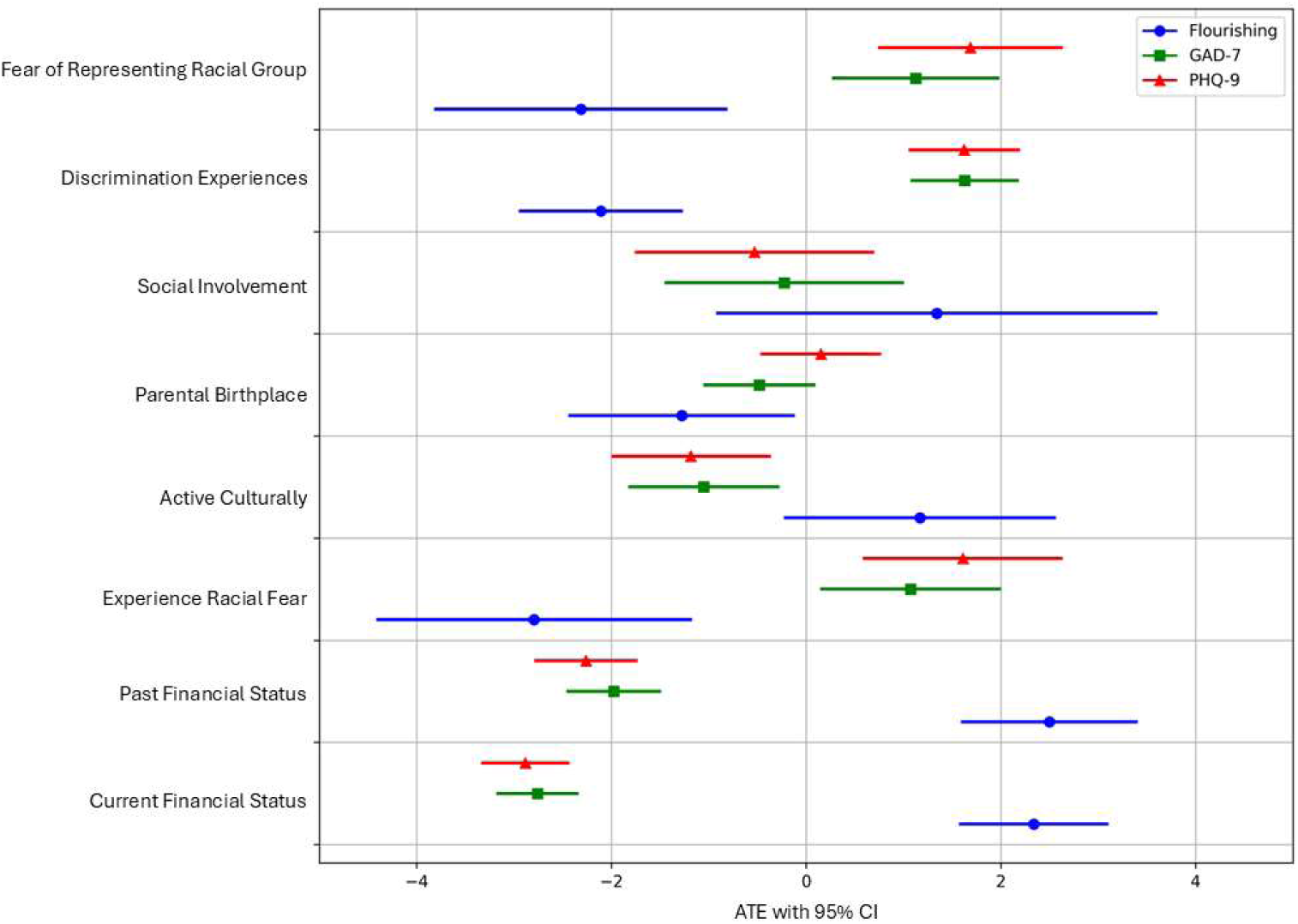
Forest plot of ATE with 95% confidence intervals calculated by the linearDML model. The ATE is the estimated influence of each variable defined in Table 2 on GAD-7, PHQ-9, and flourishing scores. Each variable has three confidence intervals listed, corresponding to the variable’s ATE on Flourishing, GAD-7, and PHQ-9 scores.

Moreover, to pinpoint the most critical variables affecting clustering, we calculate the 20th and 80th percentile values for each variable identified in the previous analysis across different clusters. Experiences of discrimination, involvement in social/cultural activities, and parents’ birthplace have non-overlapping percentiles among at-risk and flourishing clusters, meaning they are very influential in determining cluster membership.

Further, nearly all variables we identified as significantly different between racial groups of different socioeconomic status (Table 2, Table S1) have strong effects on Flourishing, GAD-7, and PHQ-9 scores (Fig. 3). These effects have an outsized impact on which cluster a data point gets assigned to.

For instance, in the standard K-Means clustering solution for Black vs White, the centroid for the flourishing cluster has a flourishing score of 49.5, a PHQ-9 score of 12.3, and a GAD-7 score of 10.3, while the getting by cluster centroid has a flourishing score of 45.8, a PHQ-9 score of 14.6, and a GAD-7 score of 12.1. Discrimination experiences are shown by the ATE analysis to decrease flourishing score by −2.1, increase PHQ-9 scores by 1.6, and increase GAD-7 scores by 1.6 (Fig. 3). Therefore, a typical flourishing student who is later affected by discrimination will see their flourishing score change to 47.4, their PHQ-9 score increase to 13.9, and their GAD-7 score increase to 11.9. These new scores more closely match the getting-by cluster than the flourishing cluster. Consequently, experiences of discrimination have such a severe effect on mental health that they can move a student from the flourishing cluster into the getting-by cluster. This finding suggests that differences in clustering results between socioeconomic status groups stem from inherent differences in the experiences of racial groups rather than disparities in sample sizes.

### Practice-Inspired Three-Subgroup Solution

In response to practitioners’ need to tailor mental health treatment, we modify our clustering algorithm to choose the minimum number of centers, k = 6, that achieves three distinct subgroups. Each center is then assigned to a flourishing, getting by, or at-risk subgroup. The centers within each subgroup are then averaged to generate an overall flourishing center, getting by center, and at-risk center (Fig. 4). We choose this constraint because practitioners may benefit from having three tiers of treatment for patients (18,42). As in the *k* = 3 algorithm, the subgroup-based algorithm shows the same cost between standard k-means clustering and socially fair clustering for groups of similar socioeconomic status. Furthermore, the subgroup-based algorithm shows that using the fairness objective considerably reduces costs for minority groups when clustering groups of different socioeconomic status (Fig. S6, S7). The convergence of the fair clustering algorithm and standard K-means algorithm for socioeconomic groups of similar status both when *k* = 3 and after cluster grouping with *k* = 6 strongly suggests that socioeconomic status plays a significant role in driving clustering outcomes. This finding implies that fairness in clustering can be partially achieved when groups have more homogeneous socioeconomic conditions. However, the benefits of using a fairness objective become particularly apparent when clustering across groups with differing socioeconomic status, as evidenced by the notable cost reduction for minority groups (Fig. S6, S7). These results emphasize the importance of considering intersecting factors like socioeconomic status in fairness-driven clustering approaches.

**Figure 4.**
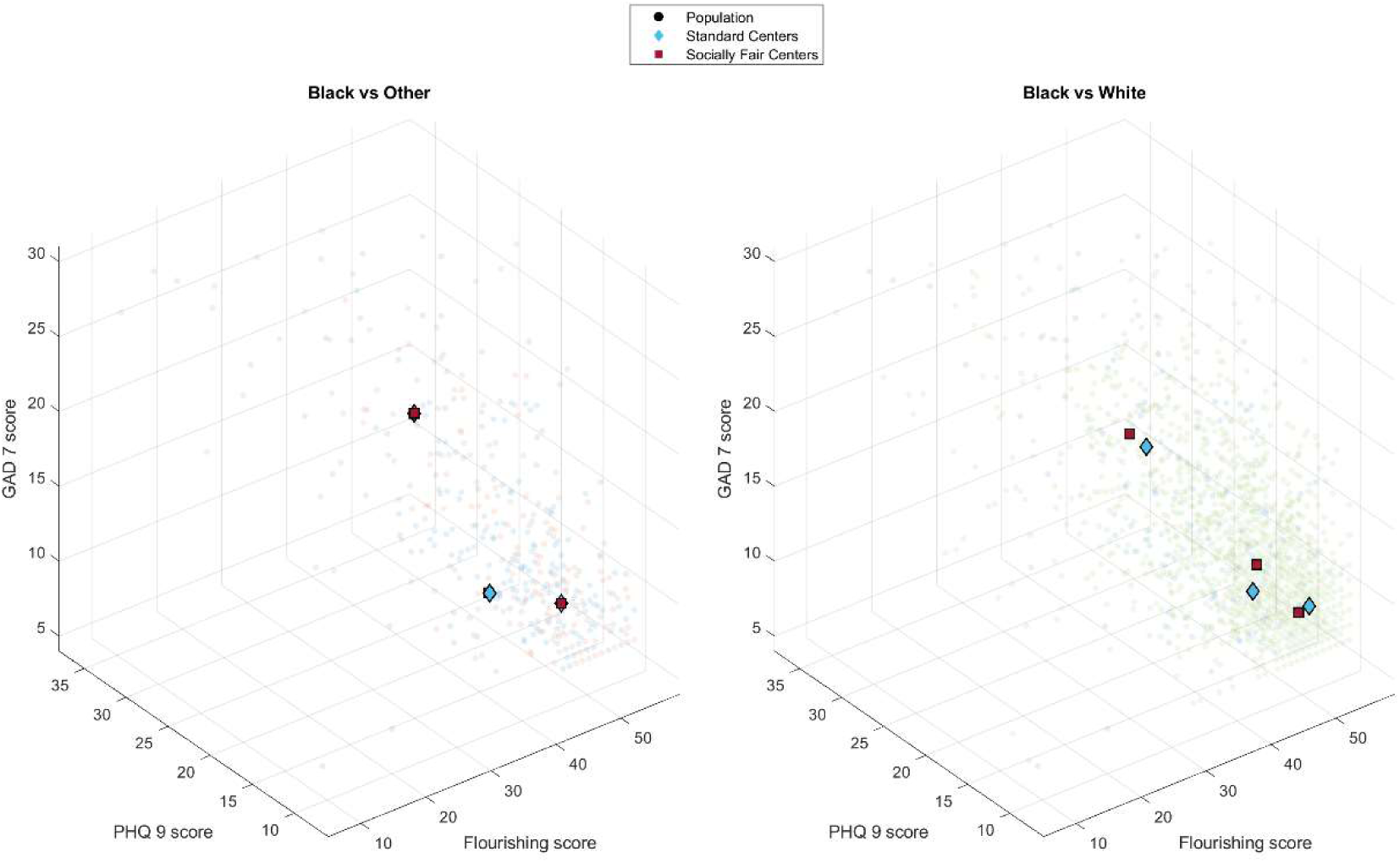
(Left) Overlay of cluster centers generated by the subgroup-based socially fair K-Means algorithm and subgroup-based standard K-Means algorithm when clustering “Black” and “Other” racial groups. (Right) Overlay of cluster centers generated by the subgroup-based socially fair K-Means algorithm and subgroup-based standard K-Means algorithm when clustering “Black” and “White” racial groups.

## Discussion

### Summary

Medical students face many mental health challenges, and the disproportionate burden experienced by racial and ethnic minority groups calls for more equitable and precise approaches to identifying those at risk. In this study, we applied a socially fair K-Means clustering algorithm to mental health data from a large, diverse sample of U.S. medical students and compared these outcomes with those derived from standard K-Means clustering. We observed that the socially fair clustering algorithm more equitably distributed the clustering costs across racial groups, reducing the cost burden for minority groups. Although minority students still had relatively higher costs than majority groups, the reduction under the fair algorithm represents a meaningful step toward more just and equitable approaches to analyzing and understanding student mental health.

We further identify that the fair clustering algorithm tends to produce a solution that approaches a two-center solution and that standard K-Means clustering also approaches a two-center solution when races share similar socio-economic backgrounds. This finding suggests that in more homogenous groups, people are either high-risk or low-risk, and decisions should be more binary. The getting-by cohort may be, in part, driven by underrepresented groups. Current models that follow a tiered approach may risk overpathologizing the middle-risk (getting by) cohorts.

### Findings and Implications

A key set of variables emerged as significantly associated with mental health outcomes, supporting the multifactorial nature of well-being in the high-pressure environment of medical training. Chief resilience factors included higher socioeconomic status, reduced perceived discrimination, financial stability, and engagement with culturally or ethnically oriented organizations. Students reporting stable current and past financial conditions consistently fared better across measures of depression, anxiety, and flourishing, supporting long-standing evidence that economic security can buffer against psychological distress. In contrast, financial strain has been linked to heightened stress, reduced access to mental health resources, and difficulty balancing the demands of medical school (47). Experiences of discrimination and the fear of negatively representing one’s racial or ethnic group had a pronounced impact on mental health. Such concerns can amplify stress, hinder class participation, and contribute to a sense of isolation. These findings are aligned with literature showing that experiences of discrimination erode well-being and academic engagement (48). Importantly, cultural or racial organizations emerged as protective spaces: students involved in these groups reported healthier mental health outcomes, possibly due to the sense of community, identity validation, and peer support these groups provide. Normalizing discussions about mental health and help-seeking behaviors also appears critical. When students feel comfortable talking about their mental well-being and accessing support services, they are more likely to seek timely interventions. Reducing stigma and creating trusted pathways to care can foster resilience and lessen the burden of mental health challenges that medical students so commonly face. These insights offer actionable pathways for medical school administrators aiming to create more equitable and supportive environments.

Addressing financial stress emerged as highly important. Schools could expand need-based financial aid, offer targeted micro-loans for emergencies, and provide financial literacy resources to help students manage debt and expenses more effectively. Such interventions could mitigate chronic financial worries and improve mental health outcomes. Clear anti-discrimination policies, supported by confidential reporting mechanisms and responsive mediation, can reduce fear and encourage students from underrepresented backgrounds to engage fully in their education. Funding and empowering culturally or ethnically oriented student organizations is another tangible step. By supporting these groups, administrators create community hubs where students can find belonging, share experiences, and collectively navigate the stressors of medical training and discrimination.

Equally important is normalizing mental health support and dialogue. Establishing regular mental health check-ins, accessible counseling services, peer support groups, and teletherapy options can help identify struggling students early, connect them to resources, and reduce the stigma that often hampers help-seeking. Ongoing evaluations through feedback surveys, focus groups, and longitudinal outcome measures ensure that interventions are effective and evolve to meet the emerging needs of minoritized students.

### Future work

We acknowledge that our study relies on voluntary self-reported data from the Healthy Minds Survey, which may introduce response bias and impact the generalizability of our findings. Furthermore, though the dataset is extensive and encompasses multiple institutions, our sample may not fully reflect the broader population of medical students across various regions and demographic groups. To better capture the experiences of students who are underrepresented in medicine, more comprehensive and inclusive data collection methods are needed. Lastly, our analysis is cross-sectional, providing a snapshot of students’ mental health at one point in time, without accounting for how these factors may change throughout their training. Longitudinal studies that include biometric data collected from wearable sensors may help bridge this gap.

### Conclusions

The current study continues the line of work defining precision well-being in medical education. Our approach refines the previously identified well-being profiles by ensuring a more equitable distribution of clustering costs across racial groups (18). The integration of socially fair clustering approaches is an important step in promoting equity within precision well-being frameworks for medical education, enabling a more nuanced understanding of student well-being. Insights from fair machine learning algorithms can guide institutional policy changes, resource allocations, and the development of inclusive and culturally sensitive support systems. Applying fairness in data-driven approaches can help medical schools create more resilient and equitable learning environments.

## Abbreviations

PHQ-9: Patient Health Questionnaire-9:
GAD-7: General Anxiety Disorder-7:
ATE: Average Treatment Effect:
DML: Double Machine Learning

## Data availability

The complete data set and code used in our analyses are available on GitHub https://github.com/AlluriPriyanshu/SociallyFairKMeans.git for review and reproducibility. The study used only openly available data by request through: https://healthymindsnetwork.org/research/data-for-researchers/

## Acknowledgments

We thank all the members of Dartmouth’s Data, Statistics, and Optimization Research Group for their comments and suggestions during the development of this manuscript. This research was supported by Dartmouth’s Class of ’74 Health Equity Scholars Program and the American Medical Association ChangeMedEd Innovation Grant Program.

## Author contributions

P.A., W.M., and T.T. designed the research; P.A. performed clustering, fairness analyses, and data preparation. P.A. and Z.C. conducted ATE analysis.; P.A., W.M., Z.C., T.T., and N. J. wrote the manuscript. W.M. and T.T. acquired the funding and provided overall supervision.

## Supplementary Material

**Figure S1.**
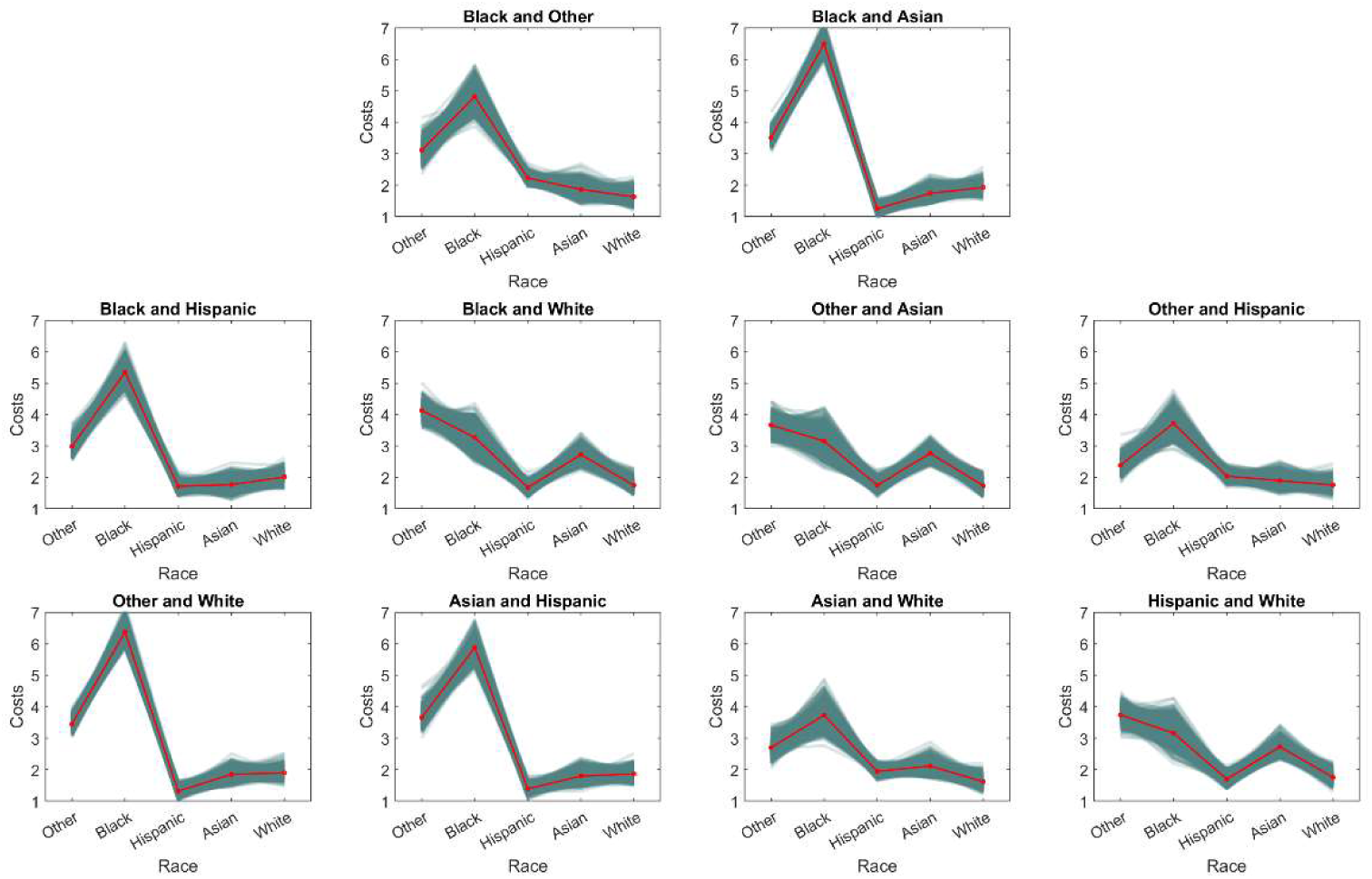
Average clustering costs across races from 10,000 bootstrap samples in the standard K-Means algorithm with three centers.

**Figure S2.**
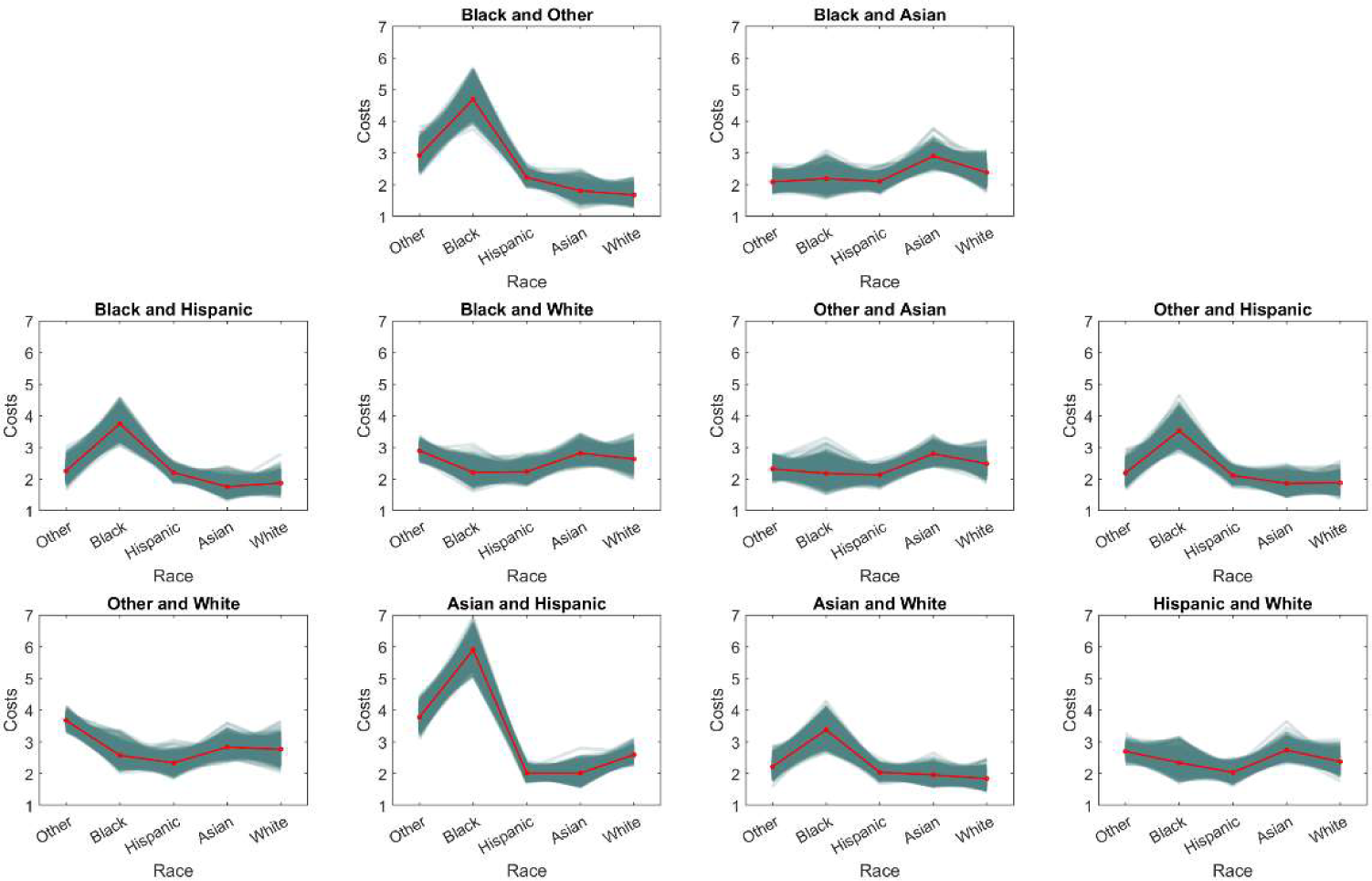
Average clustering costs across races from 10,000 bootstrap samples in the socially fair K-Means algorithm with three centers.

**Figure S3.**
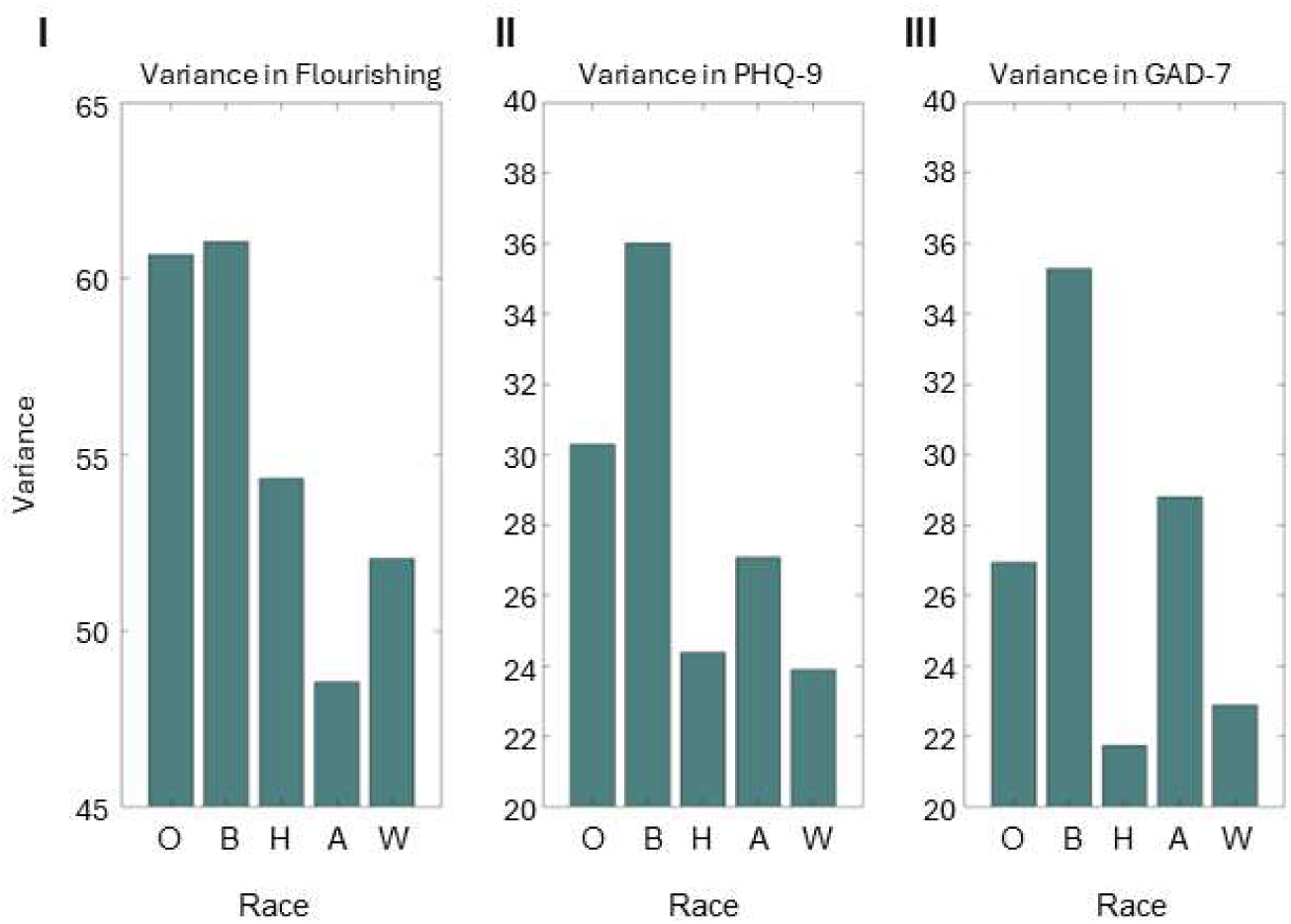
Variance in (I) flourishing scores, (II) PHQ-9 scores, and (III) GAD-7 scores across Other (O), Black (B), Hispanic (H), Asian (A), and White (W) races.

**Figure S4.**
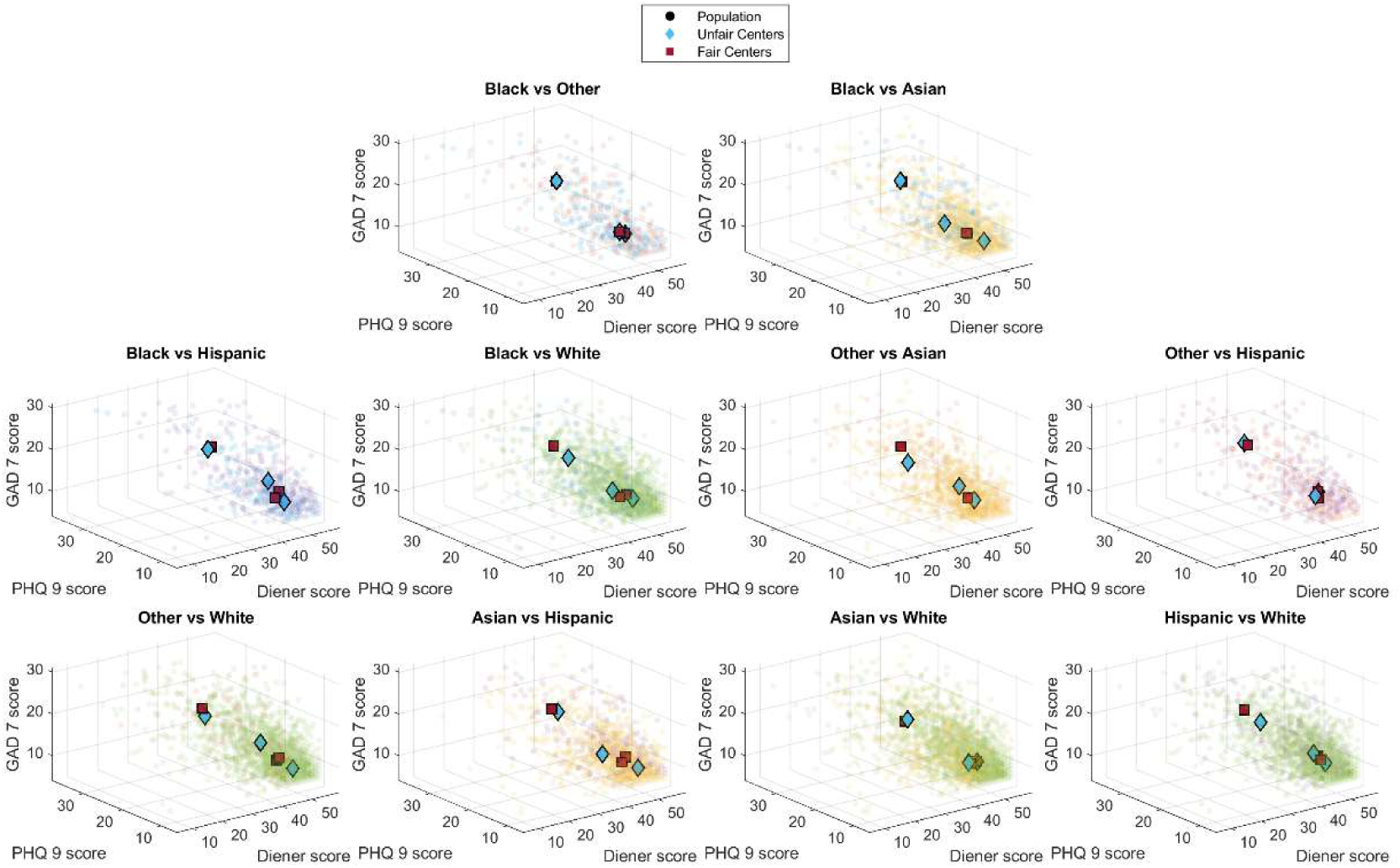
Overlay of cluster centers generated by the socially fair K-Means algorithm and standard K-Means algorithm when clustering pairs of racial groups at k = 3.

**Figure S5.**
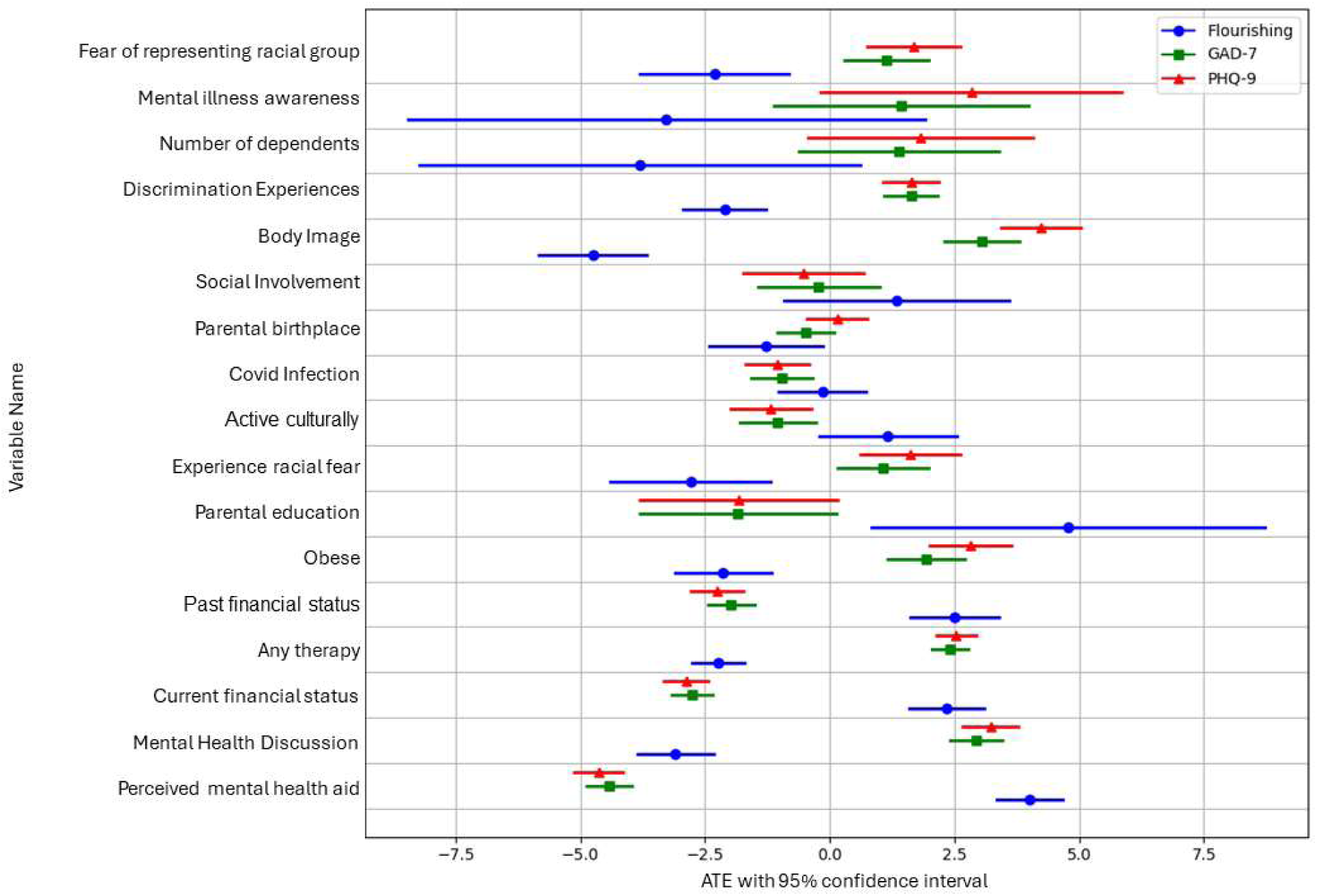
Forest plot of ATE with 95% confidence intervals calculated by the linearDML model. Variables identified as statistically significant in explaining the convergence to a two-cluster solution are plotted. Each variable has three confidence intervals listed, corresponding to the variable’s ATE on Flourishing, GAD-7, and PHQ-9 scores.

**Figure S6.**
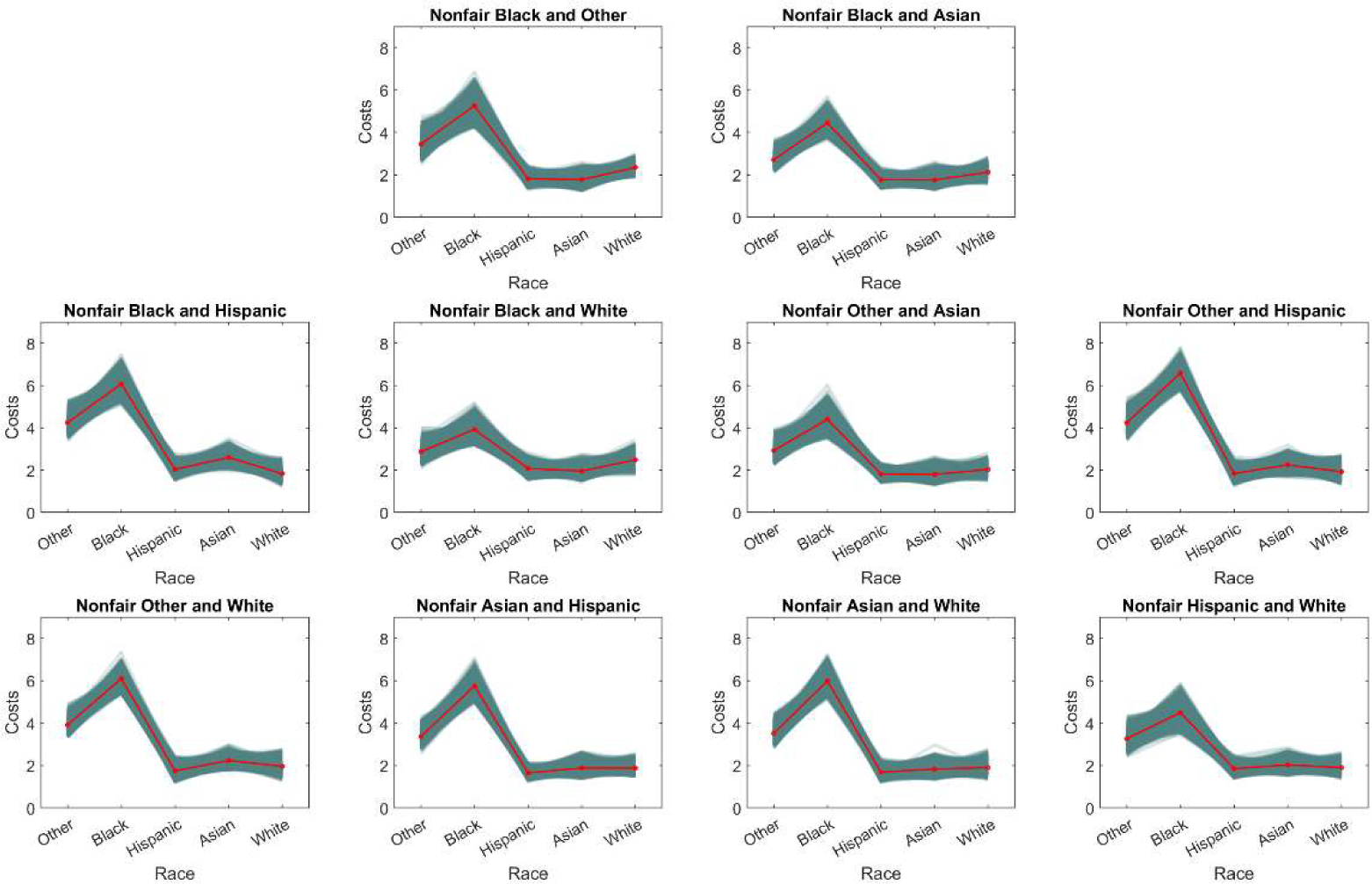
Average clustering costs across races from 10,000 bootstrap samples in the subgroup-based standard K-Means algorithm with six centers.

**Figure S7.**
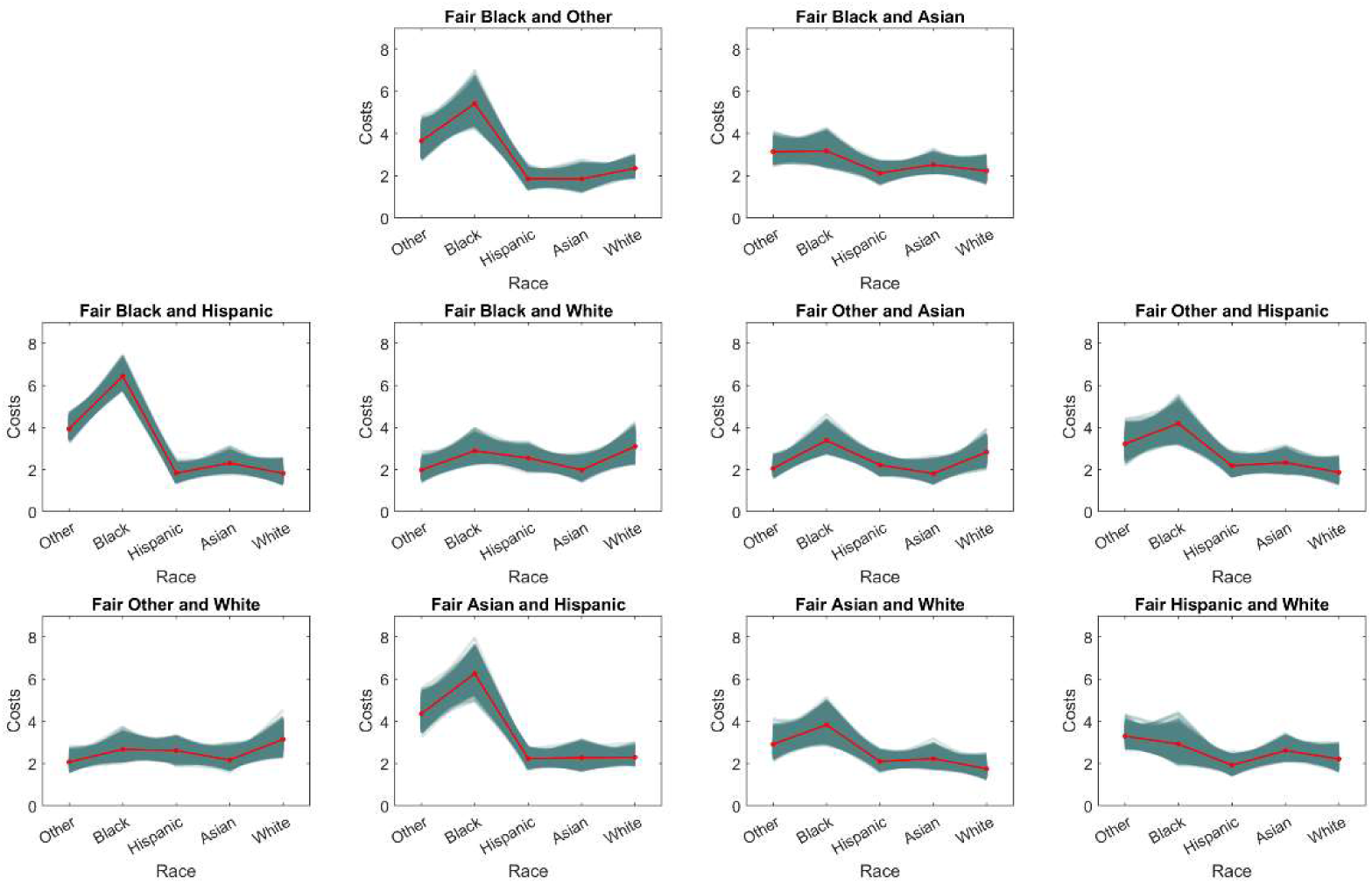
Average clustering costs across races from 10,000 bootstrap samples in the subgroup-based socially fair K-Means algorithm with six centers.

## Supplemental Tables

**Table S1.**
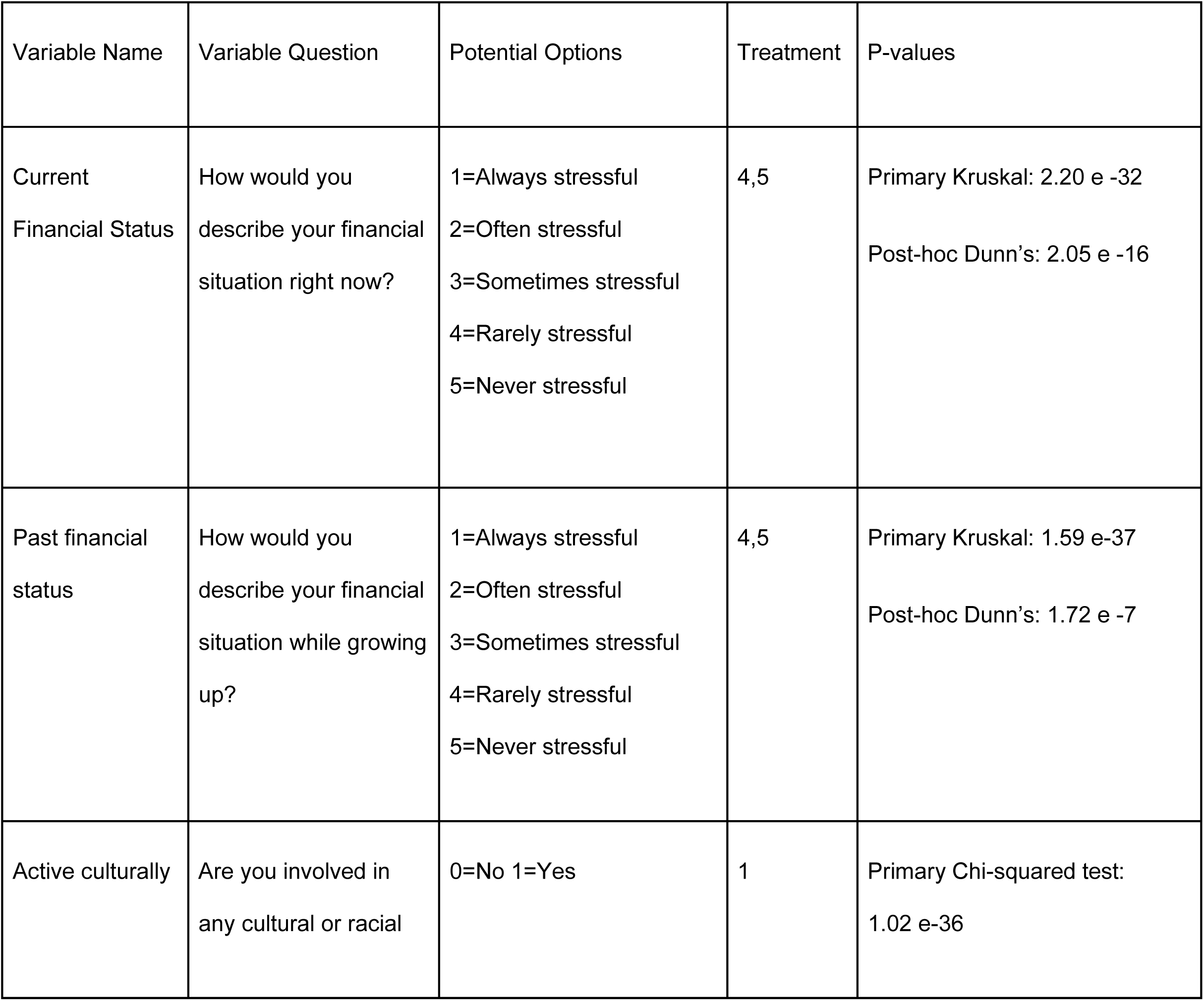
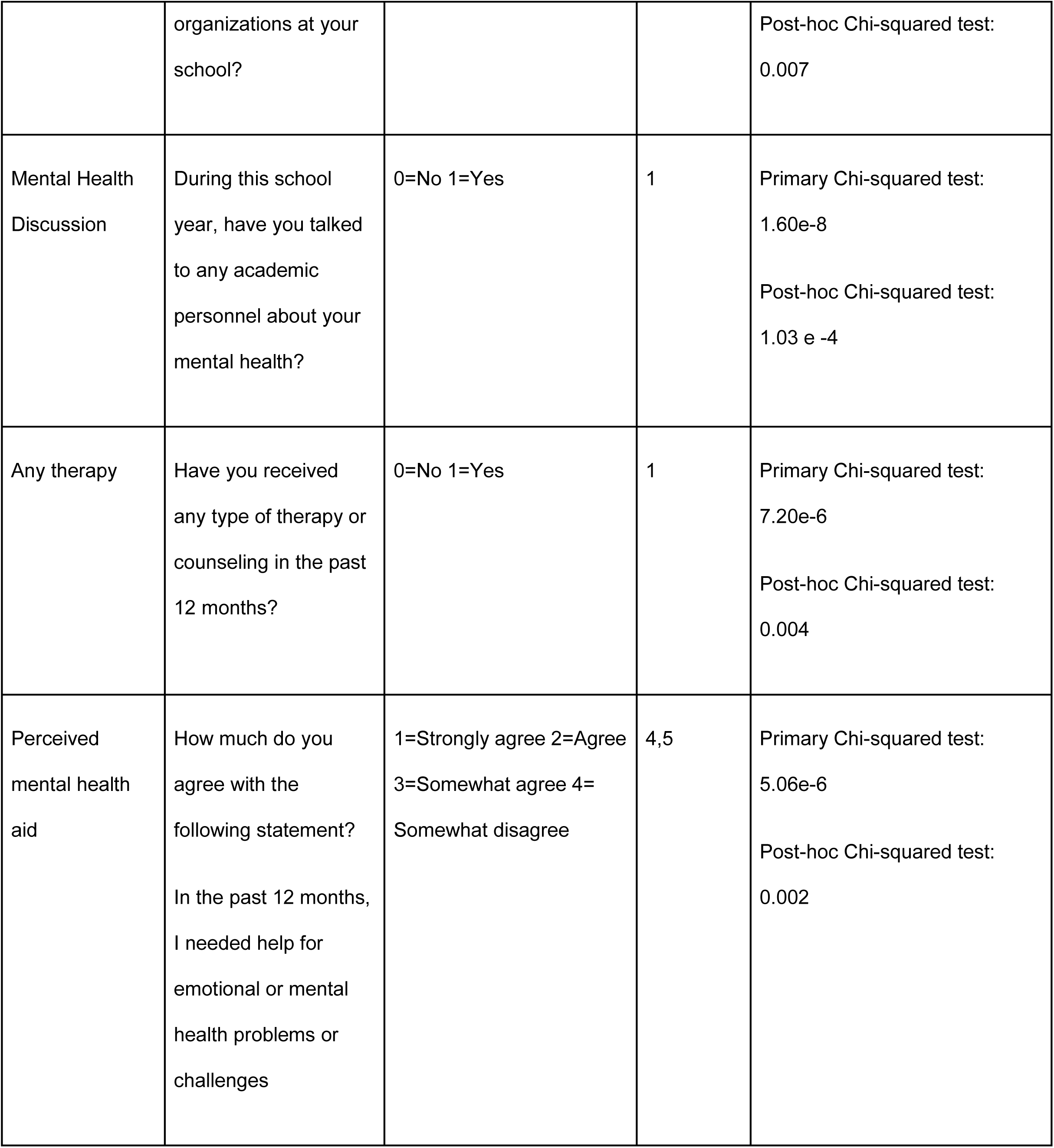
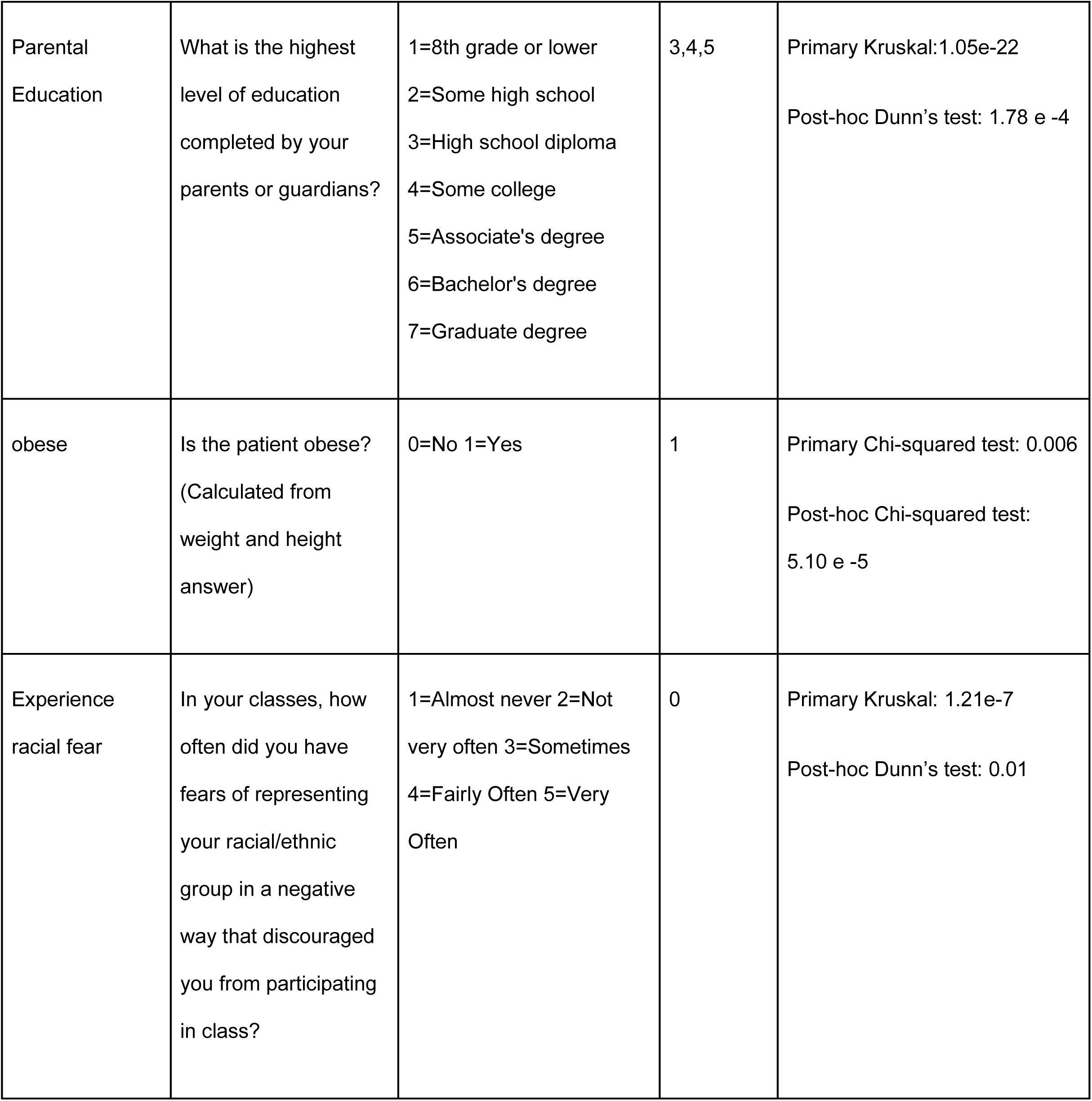
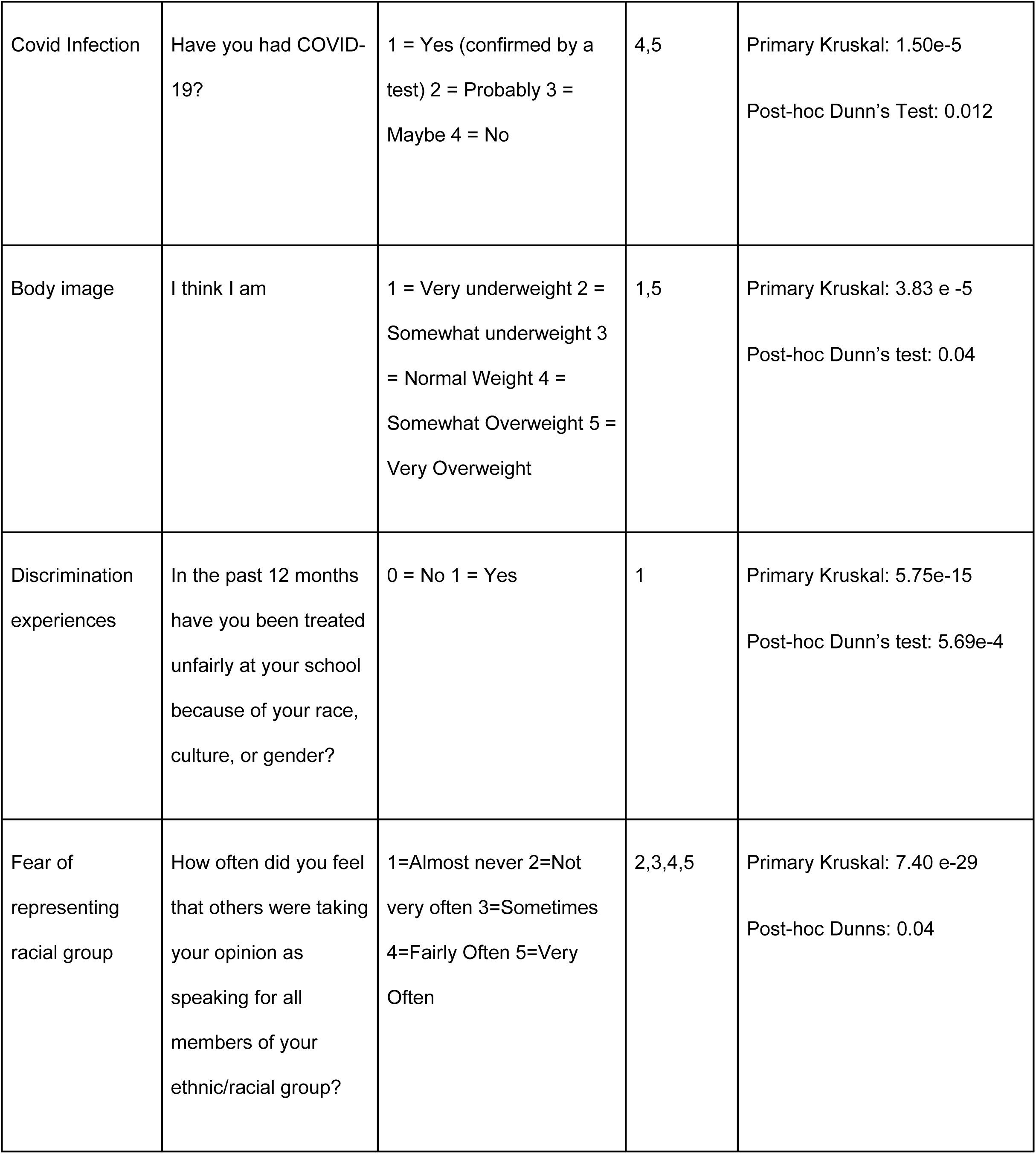
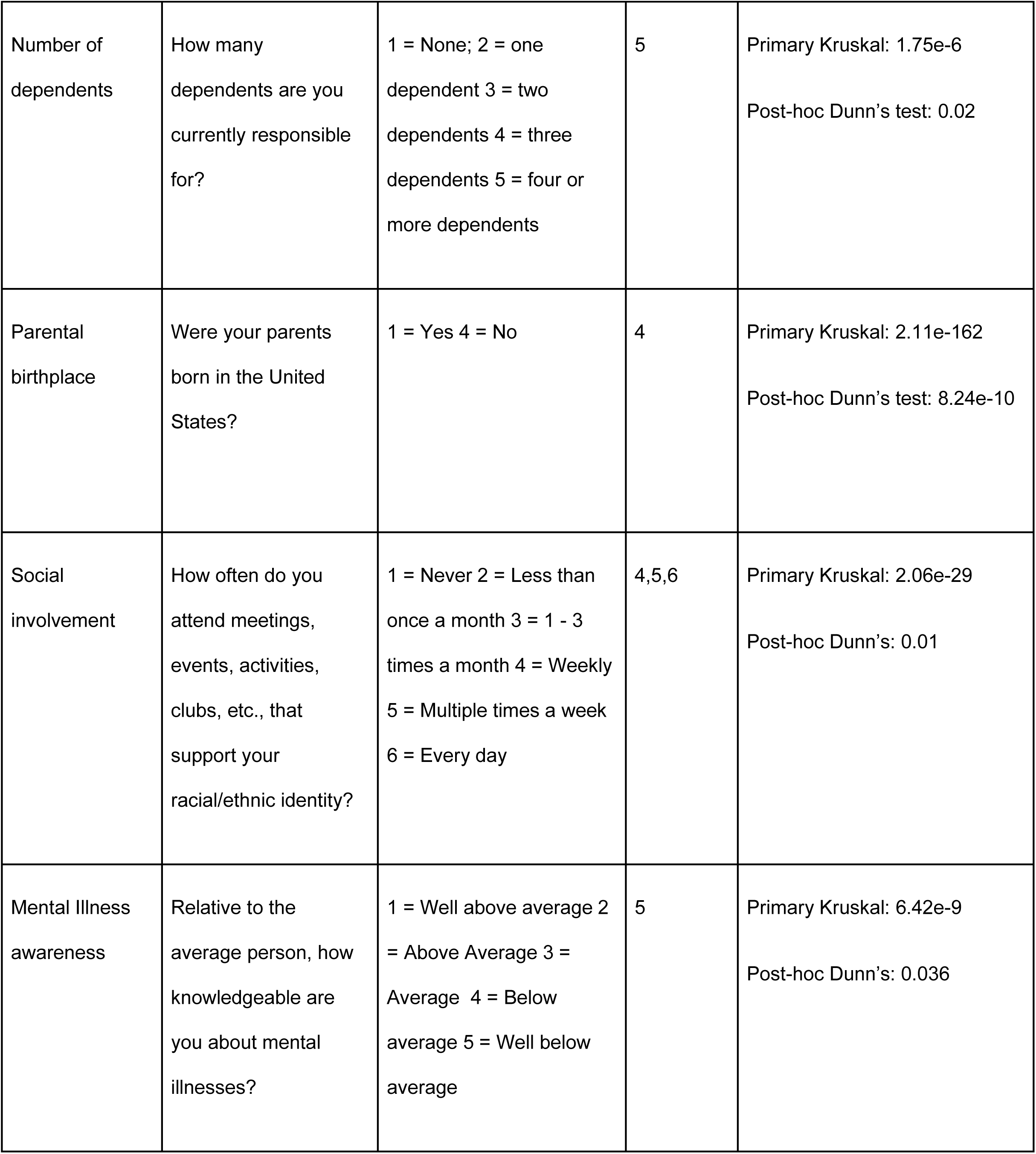
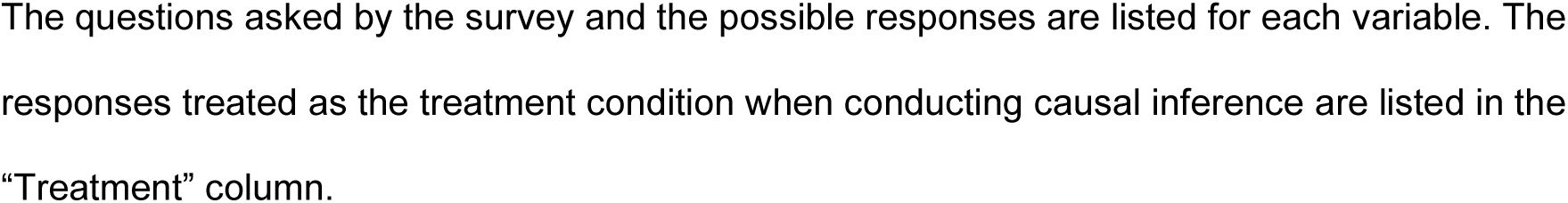
Statistically significant variables that drive differences in clustering results across groups.

| Variable Name | Variable Question | Potential Options | Treatment | P-values |
| --- | --- | --- | --- | --- |
| Current Financial Status | How would you describe your financial situation right now? | 1=Always stressful<br>2=Often stressful<br>3=Sometimes stressful<br>4=Rarely stressful<br>5=Never stressful | 4,5 | Primary Kruskal: 2.20 e -32<br>Post-hoc Dunn's: 2.05 e -16 |
| Past financial status | How would you describe your financial situation while growing up? | 1=Always stressful<br>2=Often stressful<br>3=Sometimes stressful<br>4=Rarely stressful<br>5=Never stressful | 4,5 | Primary Kruskal: 1.59 e-37<br>Post-hoc Dunn's: 1.72 e -7 |
| Active culturally | Are you involved in any cultural or racial | 0=No 1=Yes | 1 | Primary Chi-squared test:<br>1.02 e-36 |
|  | organizations at your school? |  |  | Post-hoc Chi-squared test:<br>0.007 |
| Mental Health Discussion | During this school year, have you talked to any academic personnel about your mental health? | 0=No 1=Yes | 1 | Primary Chi-squared test:<br>1.60e-8<br>Post-hoc Chi-squared test:<br>1.03 e -4 |
| Any therapy | Have you received any type of therapy or counseling in the past 12 months? | 0=No 1=Yes | 1 | Primary Chi-squared test:<br>7.20e-6<br>Post-hoc Chi-squared test:<br>0.004 |
| Perceived mental health aid | How much do you agree with the following statement?<br>In the past 12 months, I needed help for emotional or mental health problems or challenges | 1=Strongly agree 2=Agree<br>3=Somewhat agree 4=Somewhat disagree | 4,5 | Primary Chi-squared test:<br>5.06e-6<br>Post-hoc Chi-squared test:<br>0.002 |
| Parental Education | What is the highest level of education completed by your parents or guardians? | 1=8th grade or lower<br>2=Some high school<br>3=High school diploma<br>4=Some college<br>5=Associate's degree<br>6=Bachelor's degree<br>7=Graduate degree | 3,4,5 | Primary Kruskal: 1.05e-22<br>Post-hoc Dunn's test: 1.78 e -4 |
| obese | Is the patient obese?<br>(Calculated from weight and height answer) | 0=No 1=Yes | 1 | Primary Chi-squared test: 0.006<br>Post-hoc Chi-squared test:<br>5.10 e -5 |
| Experience racial fear | In your classes, how often did you have fears of representing your racial/ethnic group in a negative way that discouraged you from participating in class? | 1=Almost never 2=Not very often 3=Sometimes 4=Fairly Often 5=Very Often | 0 | Primary Kruskal: 1.21e-7<br>Post-hoc Dunn's test: 0.01 |
| Covid Infection | Have you had COVID-19? | 1 = Yes (confirmed by a test) 2 = Probably 3 = Maybe 4 = No | 4,5 | Primary Kruskal: 1.50e-5<br>Post-hoc Dunn's Test: 0.012 |
| Body image | I think I am | 1 = Very underweight 2 = Somewhat underweight 3 = Normal Weight 4 = Somewhat Overweight 5 = Very Overweight | 1,5 | Primary Kruskal: 3.83 e -5<br>Post-hoc Dunn's test: 0.04 |
| Discrimination experiences | In the past 12 months have you been treated unfairly at your school because of your race, culture, or gender? | 0 = No 1 = Yes | 1 | Primary Kruskal: 5.75e-15<br>Post-hoc Dunn's test: 5.69e-4 |
| Fear of representing racial group | How often did you feel that others were taking your opinion as speaking for all members of your ethnic/racial group? | 1=Almost never 2=Not very often 3=Sometimes 4=Fairly Often 5=Very Often | 2,3,4,5 | Primary Kruskal: 7.40 e-29<br>Post-hoc Dunns: 0.04 |
| Number of dependents | How many dependents are you currently responsible for? | 1 = None; 2 = one dependent 3 = two dependents 4 = three dependents 5 = four or more dependents | 5 | Primary Kruskal: 1.75e-6<br>Post-hoc Dunn's test: 0.02 |
| Parental birthplace | Were your parents born in the United States? | 1 = Yes 4 = No | 4 | Primary Kruskal: 2.11e-162<br>Post-hoc Dunn's test: 8.24e-10 |
| Social involvement | How often do you attend meetings, events, activities, clubs, etc., that support your racial/ethnic identity? | 1 = Never 2 = Less than once a month 3 = 1 - 3 times a month 4 = Weekly 5 = Multiple times a week 6 = Every day | 4,5,6 | Primary Kruskal: 2.06e-29<br>Post-hoc Dunn's: 0.01 |
| Mental Illness awareness | Relative to the average person, how knowledgeable are you about mental illnesses? | 1 = Well above average 2 = Above Average 3 = Average 4 = Below average 5 = Well below average | 5 | Primary Kruskal: 6.42e-9<br>Post-hoc Dunn's: 0.036 |

The questions asked by the survey and the possible responses are listed for each variable. The responses treated as the treatment condition when conducting causal inference are listed in the "Treatment" column.

